# Nocturnal systolic blood pressure dipping among people living with HIV and people without HIV: a cross-sectional study in Rural Uganda

**DOI:** 10.1101/2024.07.10.24310246

**Authors:** Isaac Sekitoleko, Sheila Kansiime, Viola Mugamba, Ismael Kawooya, Kauthra Ntabadde, Rena Nakyeyune, Femke Bannink, Moffat Nyirenda, Anxious J Niwaha, James Brian Byrd

**Affiliations:** NCD Theme, MRC/UVRI and LSHTM Uganda Research Unit, Entebbe Uganda; College of Health Sciences, Makerere University, Kampala Uganda; YouBelong, Kampala Uganda; International Centre for Evidence in Disability, London School of Hygiene and Tropical Medicine; University of Michigan Medical School, United States

## Abstract

**Background:** In this study, we investigated sleep quality, depression and stress symptoms as hypothesized factors affecting the association between HIV status and nocturnal blood pressure dipping status in rural Uganda.

**Methods:** Individuals living with HIV (PLHIV) and people without HIV (PwoHIV) underwent 24-hour ambulatory blood pressure monitoring (ABPM) and classified as extreme dippers, dippers and non-dippers based on a percentage nocturnal drop in mean systolic and diastolic blood pressure. Ordinal logistic regression models were used to assess the effect of different exposure variables (HIV status, sleep quality and other covariates) on the outcome (dipping status).

**Results:** The median age was 45 years (IQR: 35-54) and 80% of the participants were female. 24% of PwoHIV and 16% of PLHIV were overweight, 10% of HIV negative and 3% of the HIV positive individuals were obese. Depression was prevalent in both PLHIV and PwoHIV. Additionally, poor sleep quality was more prevalent in PLHIV compared to PwoHIV (70% versus 58%, P= 0.046). The study found that 53% of participants had normal dipping, while 35.1% were non-dippers, with non-dipping being more prevalent in PwoHIV individuals (34.9% vs 29.7%, P<0.001). PLHIV had 3.6 times the odds of being extreme dippers compared to PwoHIV (OR 3.64, 95% CI: 1.40 – 9.44).

**Conclusion:** This study identified high proportions of non-dipping BP profiles among both PLHIV and PwoHIV. However, the odds of being extreme dippers were higher among PLHIV compared to PwoHIV. Further research is needed to understand the underlying mechanisms contributing to extreme dipping patterns in PLHIV.

## INTRODUCTION

High blood pressure (BP) is a major cause of death globally, accounting for more than 10 million deaths per year.^1^ However, it disproportionately affects low and middle-income countries (LMICs),^2^ imposing a huge burden on health systems that are underfunded and unprepared to deal with the rapidly increasing prevalence. Monitoring BP control is central to appropriate preventive measures and titration of medication to improve outcomes among people with high BP, but accurate regular monitoring can be challenging given that BP varies over the 24-hour period and normally decreases at night by more than 10% of the mean daytime values.^3,4^ In addition, variation in the half-life of different antihypertensive medications means that a clinic visit’s time of day relative to medication dosing can add unintended variability to clinic BP measurements.^5^ As such, readings taken at the clinic or research center normally differ from those taken outside the clinic setting.^6^ Therefore, 24-hour ambulatory BP monitoring (ABPM) offers a unique and advantageous mode of BP monitoring.^7,8^

The endogenous mechanisms underlying diurnal variation in BP are orchestrated by the circadian clock via the sympathetic nervous system (SNS) and the hypothalamus-pituitary-adrenal axis (HPA).^9–11^ Sleep is crucial in maintaining the normal BP circadian rhythm that is characterised by a physiological BP reduction of 10-20% during sleep and a sharp rise just before waking up. Disturbances in sleep including reduced duration of sleep have been linked to increased risk for high BP,^12–14^ and a non-dipping BP pattern. A non-dipping BP pattern, defined as a percentage decline in night-time BP of less than 10% from daytime BP is associated with an increased risk of CVD-related morbidity and mortality^15,16^ as compared with people with a normal nocturnal dipping pattern.

Sleep and psychological disturbances (such as stress, depression, and anxiety) are two of the common extrinsic factors thought to alter BP circadian rhythm. These factors are reported to be more prevalent in people living with HIV (PLHIV) than people without HIV (PwoHIV).^17–22^ However, it is not fully known how these factors may affect the degree of BP dipping in the context of HIV and its treatment.

In this study, we compared nocturnal BP dipping profiles among PLHIV and PwoHIV. We also assessed the effect of HIV, sleep quality, psychological and biological factors on BP dipping status measured using 24-hour ABPM among individuals in rural Uganda.

## METHODS

### Study design, period and setting

We conducted a cross-sectional study at Nakaseke general hospital and Bidabujja Health Centre between April and September 2018. Nakaseke is a rural district in Central Uganda approximately 66 km north of Kampala. Nakaseke district has 7 sub-counties, 49 parishes (323 villages) with an estimated population of 197,369.^23^ The hospital has an anti-retroviral therapy (ART) clinic, which serves over 2,300 clients, receiving between 50 to 60 clients per day originating from sub-counties within and beyond the district.

### Recruitment

#### People living with HIV

The details of recruitment have been published elsewhere.^24^ Briefly, we recruited PLHIV and PwoHIV living in Nakaseke District. PLHIV were recruited from the ART clinics of Nakaseke general hospital and Bidabujja Health Centre. PLHIV aged ≥ 18 years exposed to ART and without a missed ART visit within the past 6 months were included in the study. Pregnant or currently breast-feeding women, persons with existing mental health disorders, and those who were not willing to abstain from alcohol use within 24 hours of ABPM were excluded from the study. Sampling was based on the order of attendance at the clinic.

#### People without HIV

PwoHIV were selected from the surrounding communities of Nakaseke general hospital from which the PLHIV had been recruited. Participants with a negative HIV test within 3 months prior to the study and willing to participate in the study were recruited into the study by field workers at a home visit. Participants who were willing to take part in the study but had no documented HIV negative results taken at most 3 months prior were first referred for HIV testing services. The health worker (counsellor) randomly screened participants from the community and those who tested negative were referred to the study team for eligibility assessment.

PLHIV were enrolled into HIV care and treatment at the ART clinic. Participants with undocumented recent HIV status results and unwilling to take the HIV test were excluded from the study.

### Study procedures and outcome variables

#### Data collection

Structured questionnaires were used to collect participants’ demographic information. Well-trained nurses measured height to the nearest centimetre (cm) without shoes and weight in light clothing using a SECA Stadiometer and Portable Electronic Scales. Body mass index (BMI) was calculated as weight (kg)/height (m^2^). We obtained pertinent clinical information for PLHIV, including date of diagnosis, baseline CD4+T cell count, current CD4 count, HIV treatment, HIV RNA level (which is assessed annually for all patients at the clinic) and clinic BP from the patient charts at their respective ART clinics. Information on current antihypertensive medication use and other drugs taken was self-reported by the participant. Participants’ data was collected and uploaded to a secure REDCap database hosted on Johns Hopkins University’s servers at the end of the day by trained research nurses.

#### 24-hour ABPM

24-hour ABPM was done using a 24-hour ABPM monitor (Contec Medical Systems Co., Ltd, Qinhuangdao, China) oscillometric device with an appropriate cuff for mid-upper arm circumference (small, 20–24 cm; medium, 24–32 cm; large, 32–38 cm) as previously described.^25^ But briefly, BP monitors were fitted on a regular working day (i.e. a day during which participants would be undertaking their regular daily activities). BP was automatically measured at 20-min intervals during the daytime (07:00h – 22:00h) and 30-min intervals during the night (22:00h – 07:00h). We judged the success of each 24-hour ABPM recording using criteria promulgated in the European Society of Hypertension (ESH) practice guidelines for ABPM.^26^ This approach entailed verifying: 1) that the interval between measurements did not exceed 30 minutes, 2) at least 70% of expected number of readings obtained, 3) at least 40 readings were obtained over 24 hours, 4) no more than 2 hours were missing readings, and 5) no consecutive hours were missing readings. If ambulatory BP recordings did not meet all of these criteria, they were excluded from the final analytical dataset as a quality control measure. We summarized 24-hour BP measurements as 24-hour average, daytime, and night-time average systolic and diastolic BP.

#### Psychological dimensions assessed

Sleep quality was assessed using the standard Pittsburgh Sleep Quality Index (PSQI) questionnaire, a widely used and accepted tool that has been culturally validated in various settings including African countries.^27–30^ It has seven component scores, each exploring a different sleep feature; the sum yields a global PSQI score used to define poor sleep quality when >5. Based on the sleep duration component of the PSQI score, self-reported short sleep duration was defined as <6 h of sleep per night. The following PSQI-derived data were also analysed: increased sleep latency (>30 min), reduced sleep efficiency (<85%), sleep disturbance (sleep disturbances component score >1), and daytime dysfunction due to sleepiness (daytime dysfunction component score >1). Stress was assessed by using Perceived Stress Scale (PSS-10) which measures the degree to which people perceive their lives as stressful. The Perceived Stress Scale^31^ is a widely used tool for measuring psychological stress, with good construct validity and has been used in studies in SSA.^32–35^ It is a self-reported questionnaire that was designed to measure “the degree to which situations in one’s life are appraised as stressful.”^31^ The PSS items evaluate the degree to which individuals believe their lives have been unpredictable, uncontrollable, and overloaded during the previous month. Depression was assessed using the Center for Epidemiological Studies Depression (CES-D-10)^36^ and was defined as having a CES-D-10 total score greater than 10.

### Statistical analysis

Participants socio-demographic characteristics were summarized using the mean and standard deviation or median and interquartile ranges based on the nature of the distribution of the continuous variables whereas frequencies and proportions were used to summarize categorical variables by HIV status. The Chi-square or Fisher’s exact test were used to assess associations between each of the categorical variables and HIV status whereas Kruskal Wallis test was used to compare the medians by HIV status.

Ambulatory BP profiles were determined using standard measures: average day time BP assessed as the mean and standard deviation of the individual average daytime BPs, and the nocturnal BP was the mean and standard deviation of the individual average nocturnal BPs. Individual nocturnal BP profile was assessed by dipping status (expressed as either non-dipper [<10% night fall in mean nocturnal SBP relative to mean daytime value], dipper [≥10% night fall in mean nocturnal SBP relative to mean daytime value], or extreme dipper [> 20% night fall in mean nocturnal SBP relative to daytime value] These calculations were performed as follows: percentage drop in mean daytime SBP = 100*(1-[night time mean SBP/daytime mean SBP]).

The prevalence of non-dipping and extreme dipping BP was defined as the proportion of participants with non-dipping and extreme dipping blood pressure among the total study population with complete measurements of ambulatory blood pressure. Univariable and multivariable multinomial logistic regression models were fitted to estimate the effect of HIV on nocturnal dipping, adjusting for sex, age, and BMI as *a priori* confounders. Additionally, the association between depression and stress with non-dipping BP status was also evaluated. All analysis was done using STATA version 17.

### Ethical consideration

The procedures for all the study activities were approved by Makerere University School of Medicine, Research Ethics Committee (SOM-REC: Ref 2018-019) and Uganda National Council of Science and Technology (UNCST: Ref SS 4531). Administrative authorization was provided by the Nakaseke district health officer as well as the respective hospital and clinic heads. Informed consent was obtained from all eligible participants who signed an informed consent form, or thumb-printed for those who could not read and write. Thumbprinting was attested to by a witness’ signature. Informed consent was obtained by a person who knew the patient’s primary language and discussed the study procedures, the knowledge to be gained, and the risks and advantages of participating in the study.

## RESULTS

### Socio-demographic characteristics

Overall, this study recruited 288 individuals, including 128 PLHIV and 160 PwoHIV, all of whom were included in the analysis. A majority of the participants were female 218 (80.0%). The median age of participants was 45 years (IQR: 35-54). Among PLHIV, 20.4%, 61.1%, 15.9%, 2.7% were underweight, normal, overweight, and obese, respectively. In contrast, among PwoHIV, 4.8%, 60.7%, 24.1%, 10.3% were underweight, normal, overweight, and obese, respectively (Table 1). Compared to PwoHIV, a lower proportion of PLHIV had attained at least a secondary level of education (Secondary schooling normally begins at the age of 13 and ends at the age of 18; 16.3% vs 30.7%). More of the PwoHIV were unemployed compared to PLHIV (11.3% vs 4.7%) (**Table 1**).

**Table 1:**
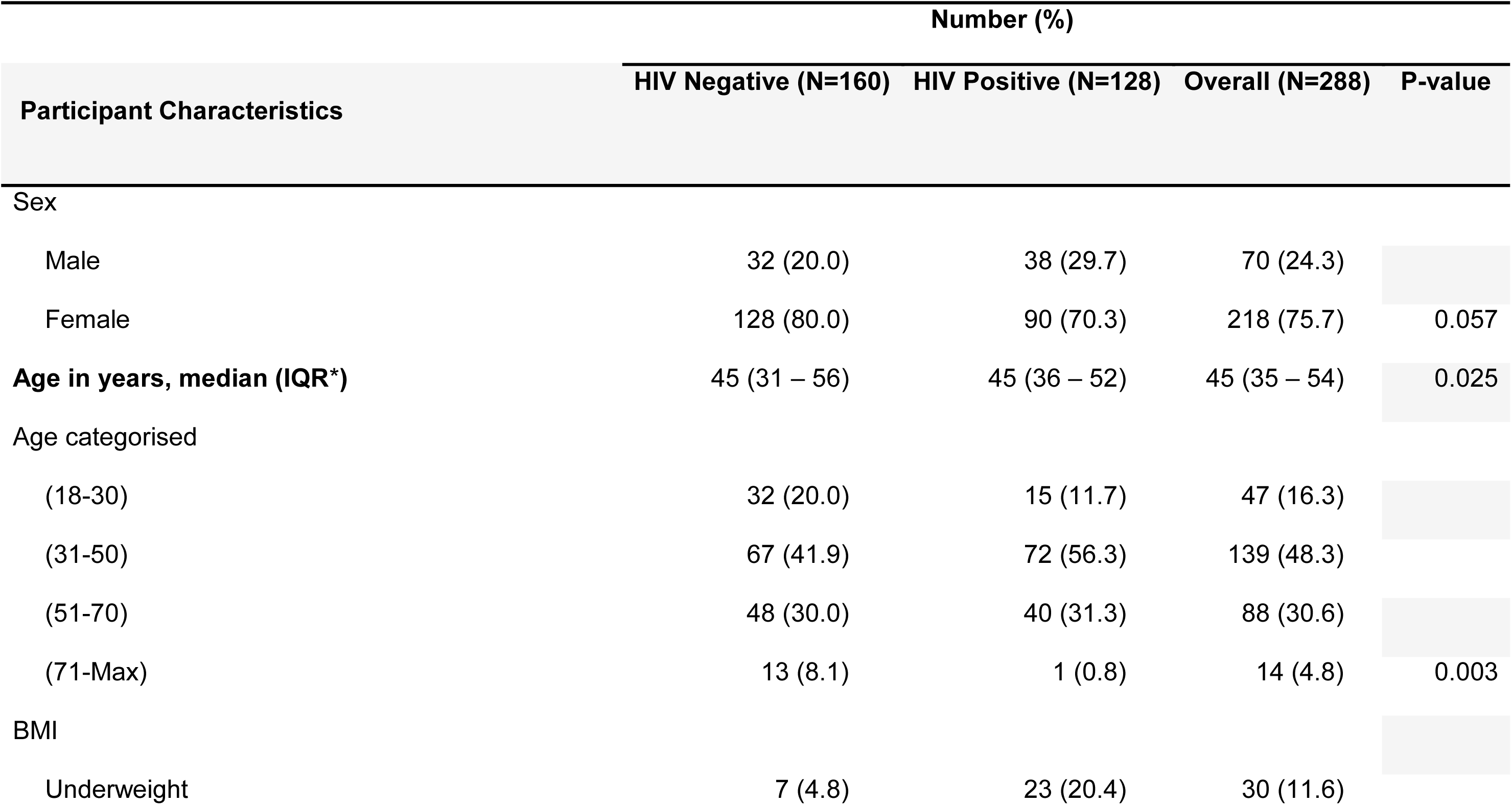
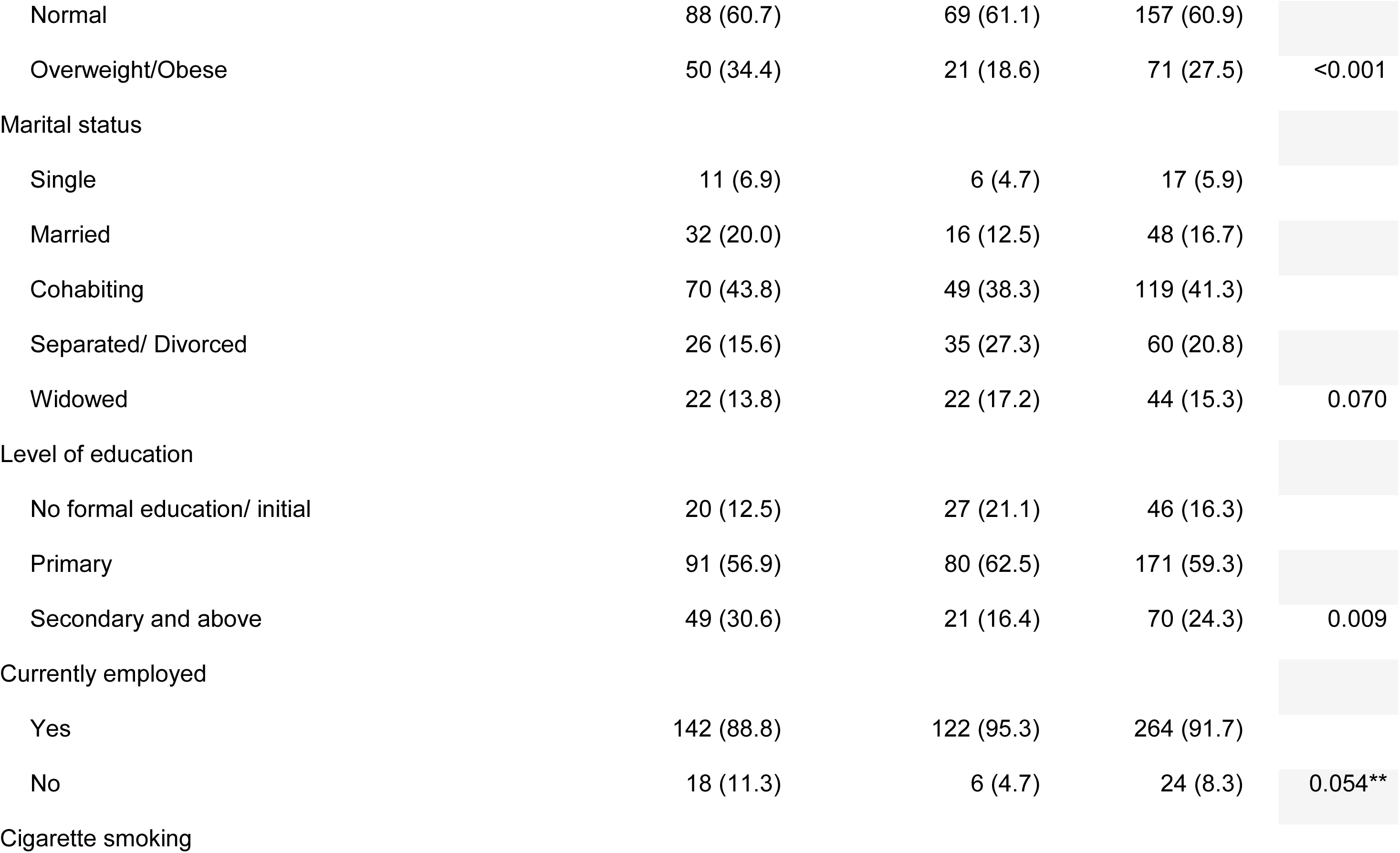
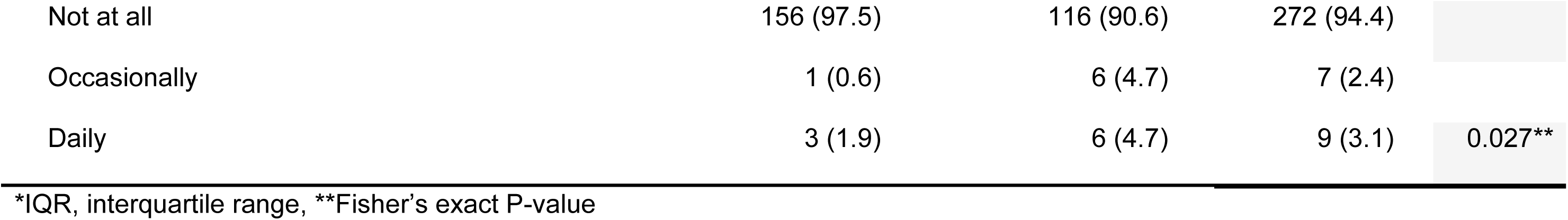
Sociodemographic characteristics of 289 individuals attending Nakaseke Hospital, by HIV status.

**Table 2:**
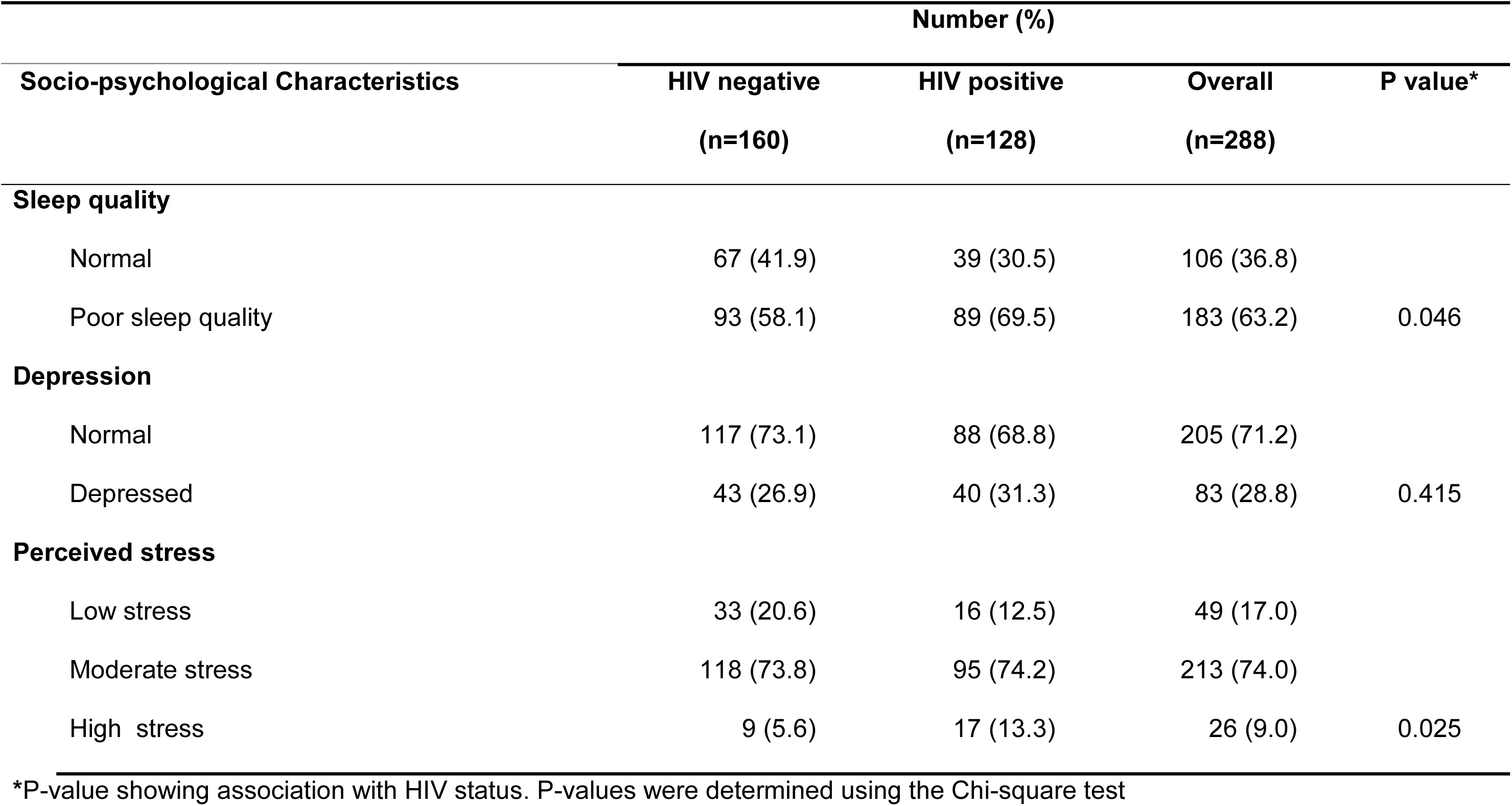
Socio-psychological characteristics of 289 individuals attending Nakaseke Hospital by HIV status.

After adjusting for HIV status, age, sex, and BMI, PLHIV had significantly lower blood pressure compared to HIV-negative patients. On the other hand, older age and higher BMI were associated with higher blood pressure (Supplementary Table 1).

### PSQI, CESD, PSS tool validation

All tools were validated for use in the study population and found to have good validity (Cronbach’s α > 0.7). The respective Cronbach’s α were as follows; PSQI: 0.73, CESD: 0.76, PSS: 0.75.

### Sleep and psychological outcomes

Overall, the proportion of participants with poor sleep quality or a high stress level was high at 183 (63.2%) and 26 (9.0%), respectively. Poor sleep quality (69.5% vs 58.1%, P=0.046) and high stress (13.3% vs 5.6%, P=0.025) were more prevalent among PLHIV (**Figure 1**).

**Figure 1.**
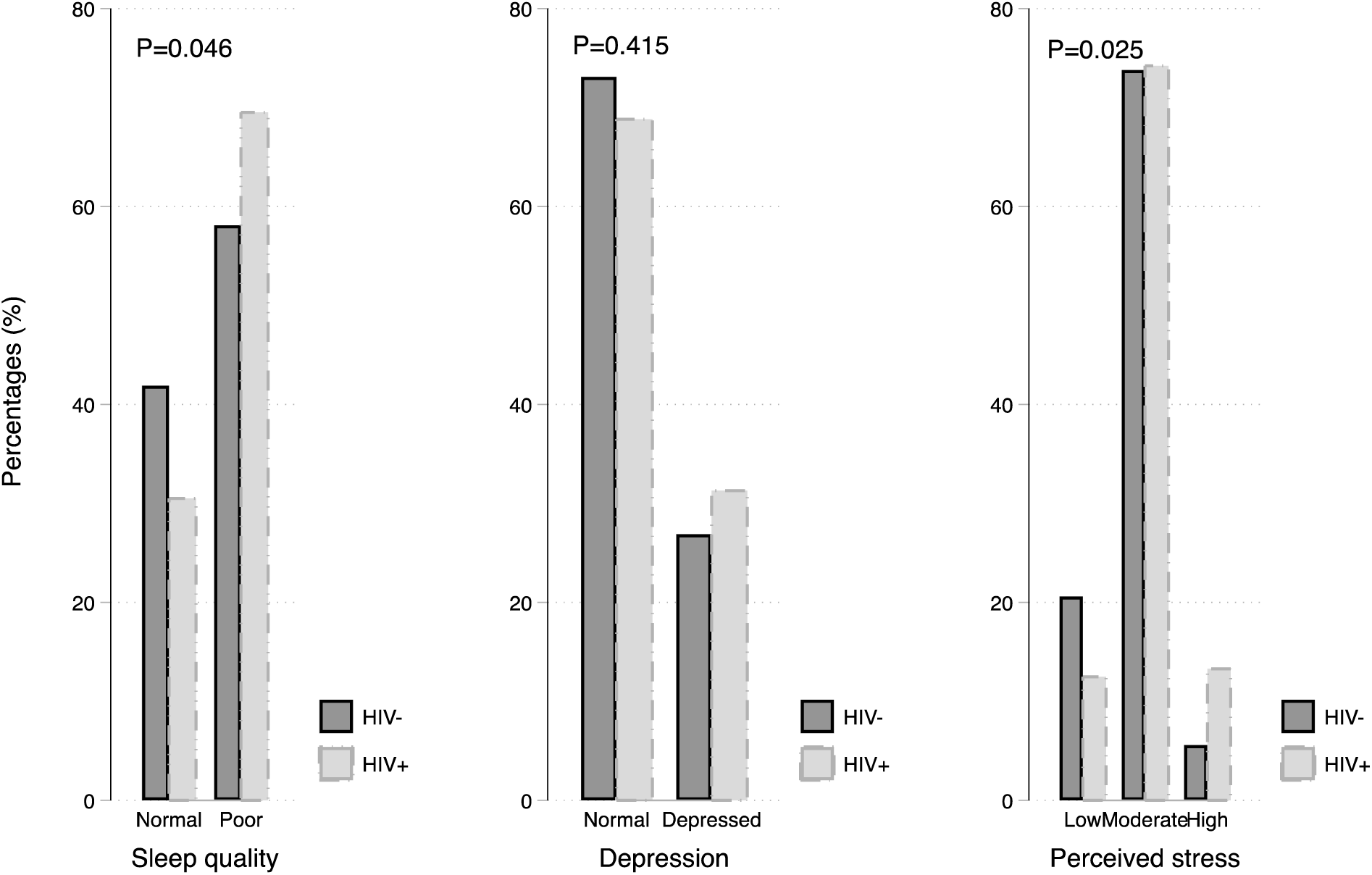
Socio-psychological characteristics of 288 individuals attending Nakaseke Hospital by HIV status.

### HIV status and systolic BP dipping

152 (53%) of the study participants had normal dipping while 101 (35.1%) were non-dippers. The prevalence of non-dipping was higher among PwoHIV (34.9% vs 29.7%), while extreme dipping was higher amongst PLHIV (20.3% vs 6.0%) (**Table 3**). Using a mean night-time BP threshold of <90/50 mmHg, a substantial proportion (25.7%) among participants with extreme dipping pattern exhibited nocturnal hypotension. This prevalence increased to 82.86% when a threshold of <100/60 mmHg was used. The prevalence of nocturnal hypertension (mean night-time BP ≥ 120/70 mmHg) within this group was considerably lower (5.7%). After adjusting for age, gender, education level, stress, and BMI, HIV was associated with extreme dipping (P<0.001). PLHIV had 3.6 times the odds of being extreme dippers compared to PwoHIV (OR 3.64, 95% CI: 1.40 – 9.44) (Table 4 and Figure 2).

**Figure 2.**
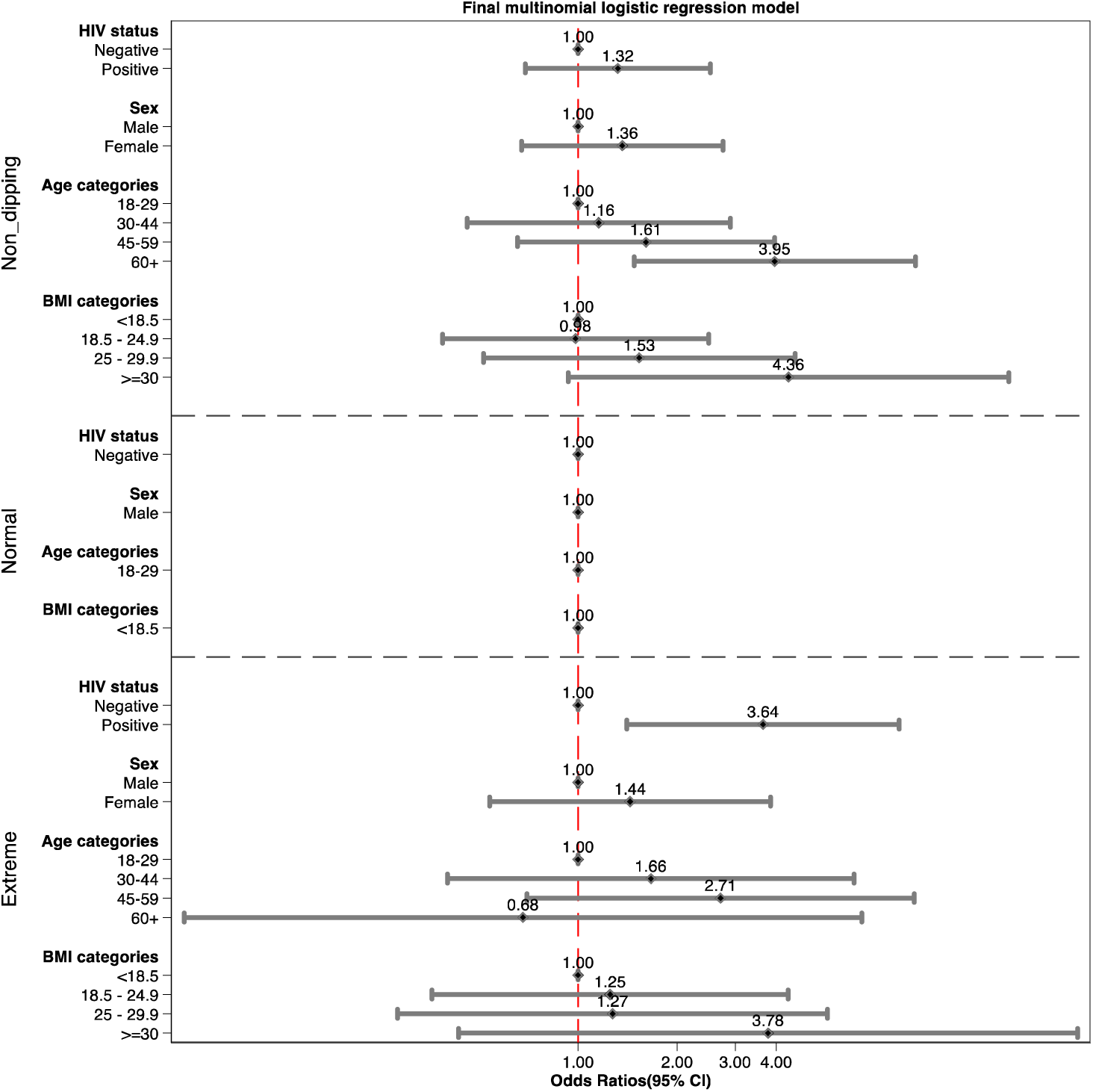
Multinomial logistic regression analysis of outcome measure systolic BP dipping.

**Table 3:** Nocturnal dipping status and median night and day systolic blood pressure among 288 HIV positive and negative.

| **Blood Pressure Feature** | **HIV Negative**<br>**(n=160)** | **HIV Positive**<br>**(n=128)** | **P value\*** |
| --- | --- | --- | --- |
| **Average SBP†, mmHg Median (IQR)** |  |  |  |
| Day-time SBP | 121.6 (114.3-131.9) | 115.7 (110.6 – 125.2) | 0.220 |
| Night-time SBP | 107.7 (99.1-120.1) | 98.8 (91.9 – 108.5) | 0.572 |
| Systolic dipping rate | 11.3 (8.0 – 14.9) | 13.8 (9.6 – 19.4) | 0.290 |
| **Dipping status n (%)** |  |  |  |
| Non-dippers | 52 (34.9) | 38 (29.7) |  |
| Normal | 88 (59.1) | 64(50.0) |  |
| Extreme dippers | 9 (6.0) | 26(20.3) | <0.001 |

\*P-values were determined using the Chi square test for categorical variables and T test for continuous values; †SBP, systolic blood pressure;

**Table 4:** Multinomial logistic regression analysis for the association between HIV status and systolic dipping.*

| Dipping Status | Variable | OR (95% CI) | P-value |
| --- | --- | --- | --- |
| Non-Dipping | HIV positive | 1.32 (0.69 – 2.52) | 0.40* |
| Normal | reference | reference | reference |
| Extreme | HIV positive | 3.64 (1.40 – 9.44) | 0.008* |

\*Adjusted for sex, age, and BMI

## DISCUSSION

In this study, we found that HIV status was associated with sleep quality as well as stress level, but not with depression. PLHIV exhibited greater proportions of poor sleep quality and elevated stress levels compared to PwoHIV. Notably, the nocturnal non-dipping BP profile was prevalent in both PLHIV and PwoHIV, with an overall prevalence of 35%. PLHIV had 3.6-fold higher odds of experiencing extreme dipping compared with PwoHIV. The high prevalence of nocturnal non-dipping and extreme dipping profiles–two of the most risky BP profiles for CVDs–underscores the need for integrating 24-ABPM in routine clinical care, including HIV care settings. ABPM predicts cardiovascular outcomes better than conventional office measurements, likely due in part to the naturalistic environment in which the measures are made outside medical settings.^7^

Our findings of high prevalence of poor sleep quality align with a recently conducted meta-analytic study, which estimated that the prevalence of sleep disturbances in PLHIV is 58%.^37^ ^38–43^ Other epidemiological studies also showed a high prevalence. Although depression was not associated with HIV in the current study, other studies have found that PLHIV often have depression and stress due to stigma and discrimination, adverse effects of medications, serious physical symptoms and financial burdens.^44^ The precise mechanisms connecting HIV and sleep disruption remain incompletely understood, and this remains an area of increasing interest. Among the plausible biological pathways are direct and indirect disturbances of sleep homeostasis induced by HIV disturbances e.g., HIV virus, ART-related effects, and chronic immune activation.^45^ ^46,47^

Although numerous studies have explored the relationship between nocturnal non-dipping and CVDs, ABPM data is still limited, particularly among PLHIV--who are at an increased CVD risk.^48^ The few studies that have explored the profiles of nocturnal blood pressure (BP) dipping among individuals living with HIV (PLHIV) in Sub-Saharan Africa (SSA) support our finding of a high prevalence of non-dipping systolic BP among PLHIV.^49,50,51^ Our observation of a prevalent non-dipping BP profile during the night among both PLHIV and PwoHIV closely aligns with recent research conducted by Nolan et al. in Tanzania.^51^ However, in contrast to our findings, Nolan et al. reported a higher, albeit statistically insignificant, prevalence of non-dipping nocturnal BP among PLHIV compared to PwoHIV (64.2% [36/56] versus 53.1% [43/81]). Notably, this disparity was particularly pronounced among participants with normal clinic BP, where two-thirds of PLHIV and only one-third of PwoHIV exhibited non-dipping nocturnal BP patterns.^51^ Furthermore, a previous systematic review and meta-analytic study, showed that compared to PwoHIV, PLHIV were more likely to have a non-dipping systolic BP pattern (OR = 2.72 [95% CI: 1.92, 3.85]).^52^

Our data showing a high prevalence of extreme dipping profile (20%) among PLHIV confirm results from the few published indicating that extreme dipping profile is prevalent, ranging from 5% to 30%.^53,54^ Within this PLHIV group of extreme dippers, more than 1 in 4 individuals experienced nocturnal hypotension. This elevated prevalence of nocturnal hypotension among PLHIV with extreme dipping profile is alarming, as it raises red flags regarding the heightened risk of cardiovascular events stemming from potential cardiac and/or cerebral hypoperfusion and subsequent organ ischemia. Indeed, there is evidence indicating that extreme dipping profile poses comparable CVD risk to non-nocturnal BP dipping.^55,56^ ^52^ Individuals with extreme nocturnal dipping--especially those with atherosclerotic arterial stenosis and excessive BP reduction from antihypertensive medication--have an elevated risk of nonfatal ischemic stroke and silent myocardial ischemia.^57^ Recent evidence suggests that extreme dipping’s association with adverse cardiovascular outcomes is strongly age-dependent, with increased risk observed above 70 years of age.^58^

The precise mechanisms leading to the heightened odds of the two abnormal BP profiles particularly among PLHIV remains unclear, further highlighting the nuanced landscape of HIV status and BP homeostasis. The proposed mechanisms for non-dipping BP include; 1) dietary salt and salt sensitivity,^59^ 2) HIV-specific factors (HIV-virus, inflammation and art),^60^ 3) Psychosocial factors, and 4) poor sleep quality. Some of the plausible mechanisms specifically for extreme dipping include: 1) HIV- and HIV proteins-induced autonomic dysfunction causing baroreflex abnormalities,^52^ 2) humoral factors such as renin-angiotensin system or insulin, 3) behavioural factors (smoking, alcohol intake, and quality of sleep),^61,62^ 4) arterial stiffness, other comorbidities and medications.^62^

We acknowledge several limitations in our study that warrant consideration. Firstly, as a cross-sectional study, it cannot establish causality regarding the effect of HIV status on BP dipping. Secondly, our use of a single 24-ABP session may not fully capture the variability inherent in blood pressure measurements, potentially leading to fluctuations in dipping status. Conducting a second ABPM measurement could mitigate this limitation. Additionally, we did not assess various factors such as arterial stiffness, inflammatory markers, hormonal profiles, dietary salt intake, and glycaemic burden, which could be associated with dipping status. Given that PLHIV exhibited a lower BMI and a higher proportion of individuals in older age groups (>70 years) compared to PwoHIV, the presence of underlying comorbidities that may influence dipping cannot be entirely ruled out. Therefore, the interpretation of results should be approached with caution, considering the potential for residual confounding that could influence the observed outcomes. This further underscores the need for further research in a larger and more diverse study population to enhance the generalizability of findings.

In conclusion, we observed high proportions of non-dipping and extreme dipping in PLHIV. The notable prevalence of extreme systolic BP dipping among PLHIV not taking antihypertensive medication underscores the critical need for further research to explore the underlying mechanisms. Additionally, more studies are needed to comprehend the influence of sleep disturbances and psychological factors on blood pressure. Investigating the potential benefits of integrating sleep therapy and psychological interventions, including counseling, into BP management is crucial for determining their effectiveness in enhancing overall BP control and preventing complications.

## Data Availability

Data will be made available upon reasonable request.

## Contribution statement

AJN, MJN, FB, IK and JBB conceptualised and designed the study. AJN researched the data. AJN, SK and IS conducted data cleaning. IS, SK and AJN analysed the data with assistance from JBB, RN and KN. IS, AJN, VM, and KN drafted the paper which was critically revised by all authors. AJN, IS, and SK had full access to all of the data in the study and took responsibility for the accuracy and the integrity of the data of analysis. All authors approved the final manuscript.

## Funding

This research project was supported by the NIH Research Training Grant (# D43 TW009340) funded by the NIH Fogarty International Center, NINDS, NIMH, and NHBLI. The content is solely the responsibility of the authors and does not necessarily represent the official views of the National Institutes of Health.

IS is also a PhD Research Fellow supported by the Fogarty International Centre of the National Institutes of Health under Award Number D43TW011401. The content is solely the responsibility of the authors and does not necessarily represent the official views of the National Institutes of Health.

## Declaration of interests

We declare no competing interests.

## Acknowledgement

We acknowledge the skilled assistance of the research assistants Kukunda Rabecca and Chemutayi Sarah, as well as the contributions of all the participants, members of staff of the Nakaseke Hospital and other health centers in Nakaseke district.

---

**Supplementary Table 1.**
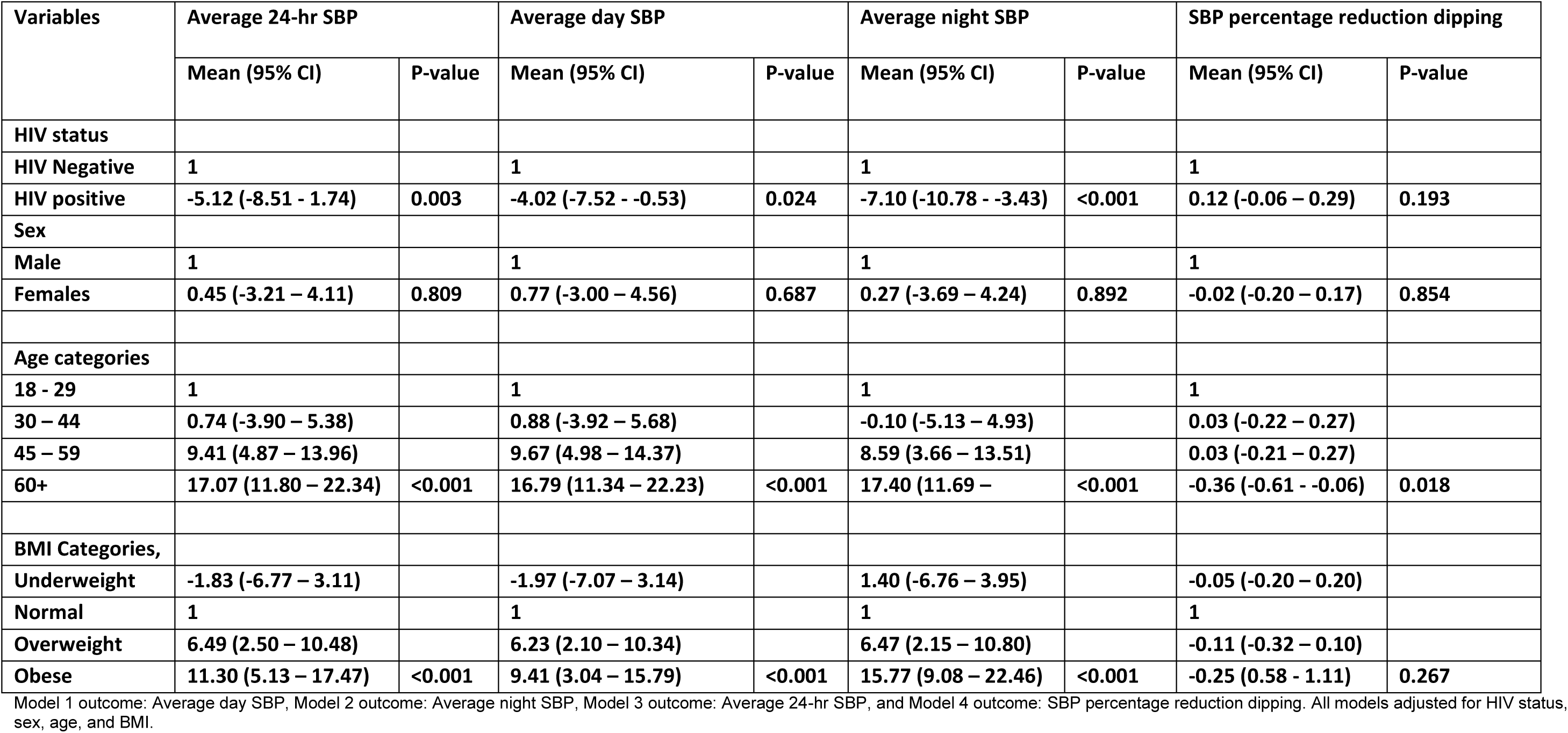
Linear Regression Models: Impact of HIV on Daytime, Nighttime, and 24-Hour Systolic Blood Pressure (SBP) and Percentage Reduction in SBP Dipping.

| Variables | Average 24-hr SBP |  | Average day SBP |  | Average night SBP |  | SBP percentage reduction dipping |  |
| --- | --- | --- | --- | --- | --- | --- | --- | --- |
|  | Mean (95% CI) | P-value | Mean (95% CI) | P-value | Mean (95% CI) | P-value | Mean (95% CI) | P-value |
| **HIV status** |  |  |  |  |  |  |  |  |
| HIV Negative | 1 |  | 1 |  | 1 |  | 1 |  |
| HIV positive | -5.12 (-8.51 - 1.74) | 0.003 | -4.02 (-7.52 - -0.53) | 0.024 | -7.10 (-10.78 - -3.43) | <0.001 | 0.12 (-0.06 – 0.29) | 0.193 |
| **Sex** |  |  |  |  |  |  |  |  |
| Male | 1 |  | 1 |  | 1 |  | 1 |  |
| Females | 0.45 (-3.21 – 4.11) | 0.809 | 0.77 (-3.00 – 4.56) | 0.687 | 0.27 (-3.69 – 4.24) | 0.892 | -0.02 (-0.20 – 0.17) | 0.854 |
| **Age categories** |  |  |  |  |  |  |  |  |
| 18 - 29 | 1 |  | 1 |  | 1 |  | 1 |  |
| 30 – 44 | 0.74 (-3.90 – 5.38) |  | 0.88 (-3.92 – 5.68) |  | -0.10 (-5.13 – 4.93) |  | 0.03 (-0.22 – 0.27) |  |
| 45 – 59 | 9.41 (4.87 – 13.96) |  | 9.67 (4.98 – 14.37) |  | 8.59 (3.66 – 13.51) |  | 0.03 (-0.21 – 0.27) |  |
| 60+ | 17.07 (11.80 – 22.34) | <0.001 | 16.79 (11.34 – 22.23) | <0.001 | 17.40 (11.69 – | <0.001 | -0.36 (-0.61 - -0.06) | 0.018 |
| **BMI Categories,** |  |  |  |  |  |  |  |  |
| Underweight | -1.83 (-6.77 – 3.11) |  | -1.97 (-7.07 – 3.14) |  | 1.40 (-6.76 – 3.95) |  | -0.05 (-0.20 – 0.20) |  |
| Normal | 1 |  | 1 |  | 1 |  | 1 |  |
| Overweight | 6.49 (2.50 – 10.48) |  | 6.23 (2.10 – 10.34) |  | 6.47 (2.15 – 10.80) |  | -0.11 (-0.32 – 0.10) |  |
| Obese | 11.30 (5.13 – 17.47) | <0.001 | 9.41 (3.04 – 15.79) | <0.001 | 15.77 (9.08 – 22.46) | <0.001 | -0.25 (0.58 - 1.11) | 0.267 |

Model 1 outcome: Average day SBP, Model 2 outcome: Average night SBP, Model 3 outcome: Average 24-hr SBP, and Model 4 outcome: SBP percentage reduction dipping. All models adjusted for HIV status, sex, age, and BMI.

## Notes

### Competing Interest Statement

The authors have declared no competing interest.

### Author Declarations

The procedures for all the study activities were approved by Makerere University School of Medicine, Research Ethics Committee (SOM-REC: Ref 2018-019) and Uganda National Council of Science and Technology (UNCST: Ref SS 4531).

